# Deep Learning-based Prediction of Early Cerebrovascular Events after Transcatheter Aortic Valve Replacement

**DOI:** 10.1101/2021.09.10.21263380

**Authors:** Taishi Okuno, Pavel Overtchouk, Masahiko Asami, Daijiro Tomii, Stefan Stortecky, Fabien Praz, Jonas Lanz, George CM Siontis, Christoph Gräni, Stephan Windecker, Thomas Pilgrim

## Abstract

**Background:** Cerebrovascular events (CVE) are one of the most feared complications of transcatheter aortic valve replacement (TAVR). CVE appear difficult to predict due to their multifactorial origin incompletely explained by clinical predictors. We aimed to build a deep learning-based predictive tool for TAVR-related CVE.

**Methods:** Integrated clinical and imaging characteristics from consecutive patients enrolled into a prospective TAVR registry were analysed. CVE comprised any strokes and transient ischemic attacks. Predictive variables were selected by recursive feature reduction to train an autoencoder predictive model. Area under the curve (AUC) represented the model’s performance to predict 30-day CVE.

**Results:** Among 2,279 patients included between 2007 and 2019, both clinical and imaging data were available in 1,492 patients. Median age was 83 years and STS score was 4.6%. Acute (<24 hours) and subacute (day 2-30) CVE occurred in 19 (1.3%) and 36 (2.4%) patients, respectively. The occurrence of CVE was associated with an increased risk of death (HR [95%CI]: 2.62 [1.82-3.78]). The constructed predictive model uses less than 107 clinical and imaging variables and has an AUC of 0.79 (0.65-0.93).

**Conclusions:** TAVR-related CVE can be estimated using a deep learning-based predictive algorithm. The model was implemented online for broad usage. (https://www.welcome.alviss.ai/#/cvecalculator).

## INTRODUCTION

Deep learning is a subset of machine learning where artificial neural networks, algorithms inspired by the structure and function of the human brain, learn complex relationships from large amounts of data to make accurate predictions^1,2^. In certain fields, artificial neural networks have been shown to exceed human abilities^1,2^. The ability of deep learning to recognize patterns and learn valuable features from raw input data without requiring human intervention has a potential to help in acquiring, interpreting, and synthesizing growing medical data to improve clinical care^3^. Thus, deep learning is now increasingly investigated and utilized in the medical filed, of which cardiovascular medicine is no exception^4,5^. As a wide variety of data such as clinical data, laboratory data, imaging data, and procedural data needs to be integrated and considered for preprocedural planning, performing, and post-procedural care of transcatheter aortic valve replacement (TAVR), the field may particularly benefit from the implementation of deep learning^6-8^.

Cerebrovascular events (CVE) are rare but yet the most feared complications in patients undergoing TAVR^9^. Despite continuous improvements in technique and devices, the risk of CVE remains fairly stable across randomized as well as observational studies^10,11^. Hence, the challenge of CVE prevention is an unmet clinical need and will gain particular topicality with the extension of TAVR to lower risk and younger patients at hand. Histological analysis of debris captured by cerebral protection devices during TAVR has shown that not only thrombus, but also aortic valve and wall tissue, calcium and connective tissue embolize in the cerebrovascular circulation during the procedure, suggesting multiple and complex mechanisms of CVE in patients undergoing TAVR^12^. Although several independent risk factors, such as atherosclerosis, atrial fibrillation, use of balloon dilatation, and device dislocation or embolization have been suggested^9,13-18^, no predictive tool regrouping all possible contributing factors has been proposed so far. This might be because conventional statistical approaches such as logistic regression fail to integrate the multifactorial interactive relationship of numerous possible predictors of CVE which statistically remain “rare” events.

We therefore aimed to apply a deep learning method to develop a predictive model for CVE after TAVR.

## METHODS

### Study population

Between August 2007 and February 2019, clinical, procedural and follow-up data of 2,279 consecutive patients undergoing TAVR for aortic stenosis were prospectively enrolled into an institutional registry that forms part of the Swiss TAVR registry (NCT01368250)^11^. For the purpose of the present study, we selected patients with adequate preprocedural multi-detector computed tomography (MDCT) data from the registry as we hypothesized that imaging data is important for CVE predicting modelling.

The indication for TAVR was decided based on the evaluation of the local Heart Team. The procedure was performed according to standardized protocols regarding the access site, type and size of the device, based on a comprehensive evaluation of clinical, biological and anatomical characteristics as per echocardiography and MDCT for each patient at baseline. Procedural anticoagulation was achieved with administration of intravenous heparin at an initial dose of 5000 IU or 70 IU/kg, aiming at an activated clotting time (ACT) of 250 to 300 seconds. The preferred antithrombotic treatment after TAVR comprised dual antiplatelet therapy (aspirin and clopidogrel) for 6 months followed by lifelong aspirin in patients without indication for oral anticoagulation, but ticagrelor or prasugrel were accepted in presence of concomitant indication such as recent percutaneous coronary revascularisation. In patients with atrial fibrillation or other indication for oral anticoagulation, the anti-thrombotic regimen comprised an anticoagulant agent alone or a combination with single or dual antiplatelet therapy as tailored according to patient comorbidities. The registry is approved by the Bern cantonal ethics committee and all participants provided written informed consent prior to inclusion. The study was conducted in compliance with the Declaration of Helsinki.

### Computed tomography evaluation

Contrast-enhanced ECG-gated MDCT examinations were performed on either a Siemens Somatom Sensation Cardiac 64 scanner with a slice collimation of 1.5 mm or a Siemens Somatom Definition Flash Dual-Source scanner with a slice collimation of 0.6 m, tube voltage of 100 or 120 kV, and tube current according to patient size (Siemens Medical Solutions, Inc., Forchheim, Germany) as previously described^19^. Acquired CT images were independently re-evaluated on a dedicated workstation (3mensio Structural Heart, 3mensio Medical Imaging BV, Bilthoven, The Netherlands). Basic measurements were made in accordance with a current expert consensus document^20^. Calcium volume in the aortic valve complex and mitral valve complex were measured as previously described^19,21^. Ascending aorta length is defined in the presented study as the distance at which the line drawn perpendicular to the aortic valve annulus plane hits the ascending aortic wall (**Figure 1**).

**Figure 1:**
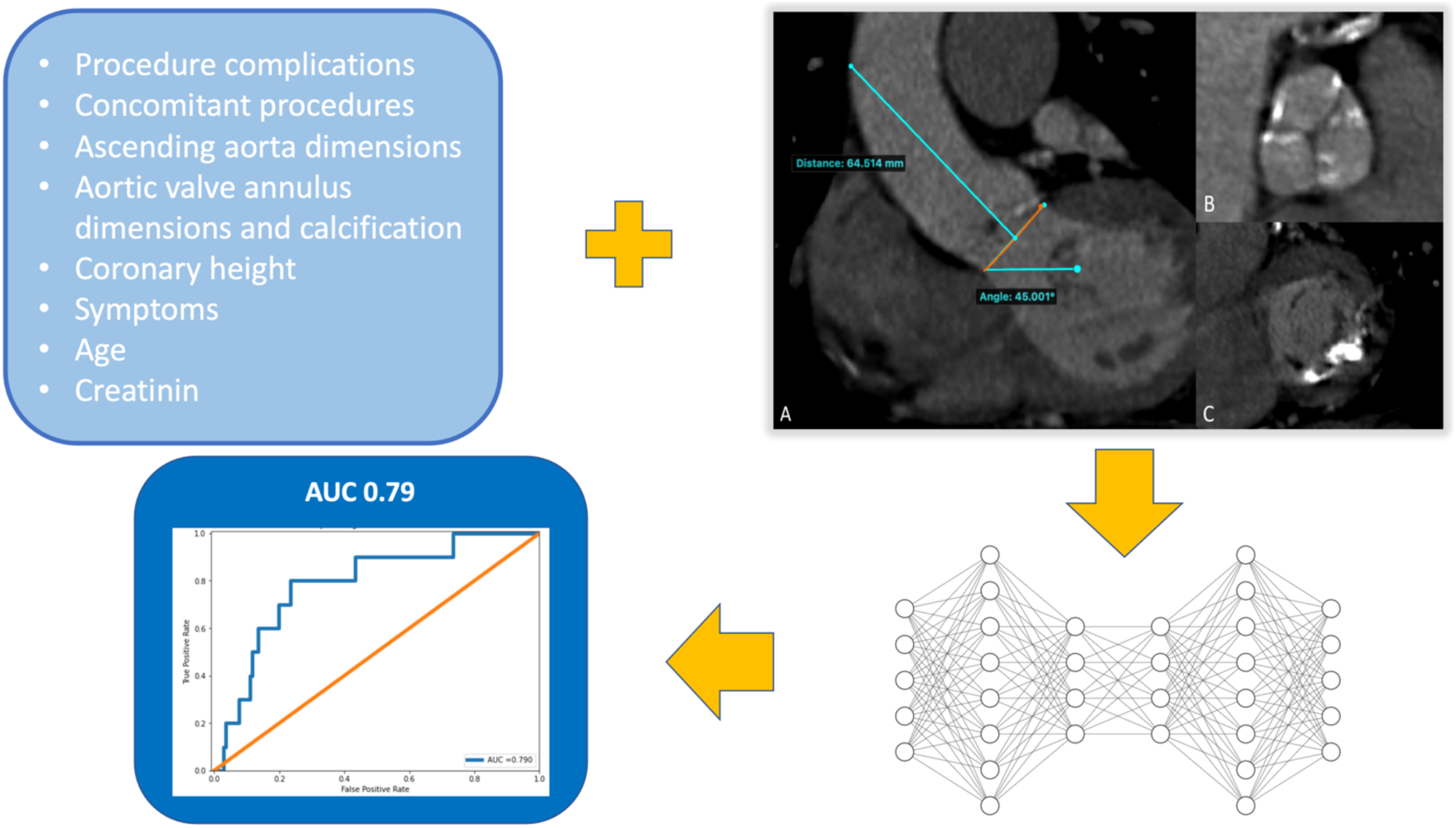
Graphic representation of the presented predictive model. CT imaging and other clinical data are entered in the autoencoder predictive model to yield an estimation of the risk of a cerebrovascular event (stroke or transient ischemic attack). A: CT sagittal view of the ascending aorta and left ventricle outflow tract. In orange is presented the aortic annulus plane. In cyan are presented the ascending aorta length and the annulus angulation. B: transverse view of aortic valve cusps with calcium in the setting of a degenerative aortic stenosis. C: transverse view of a mitral valve with calcium. Ascending aorta length: distance at which the line drawn perpendicular to the aortic valve annulus plane hits the ascending aortic wall (important note: this is different from the usual measurement which corresponds to the distance between the brachio-cephalic trunc and the aortic annulus). Right coronary cusp height: perpendicular distance between the outflow of the right coronary and the aortic annulus. LVOT: left ventricle outflow tract. The autoencoder model is represented here by a simplified schema.

### Study objectives

The primary objective of this study was to build a predictive model for TAVR-related cerebrovascular events. We also aimed to investigate the relative importance of clinical and CT imaging variables for CVE risk prediction, and to evaluate the impact of CVE on mortality.

The primary endpoint was the area under the receiver operating curve (AUC), the most widely used measure of global performance of predictive models.

### Clinical endpoints/outcomes

All data were entered into a dedicated web-based database held at the Clinical Trials Unit of the University of Bern. A clinical event committee adjudicated all adverse events based on the Valve Academic Research Consortium (VARC) criteria^22^. Vital status and date of censoring or death were recorded in the registry.

All CVE, including any stroke and transient ischemic attack (TIA), were recorded. Whenever stroke and transient ischemic attack was suspected, patients underwent examination by a board-certified neurologist and underwent diagnostic neuroimaging at his discretion. Stroke and transient ischaemic attack were defined in accordance with the definition of a central nervous system (CNS) type 1 and type 3a event, respectively (ARC)^23^. Stroke events were further subdivided into disabling and non-disabling as per VARC-2^22^. We hypothesized that TAVR related CVE occurred within 30 days after the procedures and considered all acute and subacute CVE as the primary outcome for predictive modelling.

### Deep learning predictive modelling

Machine learning modeling refers to the development of a mathematical representation of data by a training process. Given the highly unbalanced character of the predicted class (only 3.7% of patients presented a 30-day CVE versus 96.3% did not) we chose a rare-event autoencoder to be the prediction model^24^.

The rare event autoencoder is made of 2 modules of neural networks: encoder and decoder. The former learns the underlying features representing the input data while the latter tries to recreate the original data from the features learned. The model is trained using the data of patients who did not present an event. When confronted with new data a reconstruction mean squared error (MSE) between the predicted data and the ground truth was used to estimate the risk of CVE. When the MSE is above a specified threshold we consider that the patient is at risk of CVE. Clinically it corresponds to an estimation of how a given patient differs from the population that usually does not develop a 30-day CVE regarding a range of clinical, biological and imaging characteristics. The threshold is chosen from the analysis of the precision-recall curve of the validation data (**Supplementary Figure 1**).

Categorical variables were entered in the model after one-hot-encoding pre-processing. Variable importance was used for variable selection and estimated based on the impact of its neutralization (median value for continuous variables and 0 for categorical variables) on model performance. The selected variables are displayed ranked by importance in **Supplementary Table 2 and Figure 2**.

### Statistical analysis

Continuous and categorical variables are presented as medians (interquartile range) and as counts (percentage). Those were assessed with Mann–Whitney U test for continuous variables, and Chi square or Fisher exact test as appropriate for categorical variables. Collinearity was assessed prior to model building by analysis of the correlation matrix. Missing values ranged from 3.2 to 17.5% (overall 7.6%) and were treated with multiple imputations by a Monte Carlo dropout autoencoder^25^.

Model performance was evaluated on the test set calculating the AUC The overall database was randomly split into a train dataset (2/3 of total) used for model training and an independent test dataset (1/3) used for performance evaluation. The validation set represented 20% of the training set. The model was trained using python 3.7.9 and keras 2.4.3 software on MacBook Pro 2.4 GHz 8-Core Intel Core i9.

Python 3.7.9 (Python Software Foundation), R 3.6.1 (R Core Team, R A language and environment for statistical computing, R Foundation for Statistical Computing (Vienna, Austria. URL https://www.R-project.org/) and SPSS 23 (IBM SPSS Statistics for Windows, Version 23.0 Armonk, NY: IBM Corp) software were used to perform the statistical and machine learning analysis. P-value <0.05 was considered significant unless otherwise specified.

## RESULTS

Out of a total of 2279 patients included in the Bern TAVR database, 1492 patients had complete clinical and MDCT data (65%) (**Figure 2**). Baseline characteristics are summarized in **Tables 1 and 2**. The median age of the population was 83.2 years (interquartile range [IQR] 79.4-86) and 48% were female. Twelve percent of patients had a history of prior stroke or TIA, and one third of patients had a history of atrial fibrillation. After a median duration of follow-up of 3 (1-4.3) years, 105 patients had experienced at least one CVE (7.8%) during the follow-up period. CVE occurred within the first 24 hours in 19 patients (acute CVE), and between day 1 and day 30 in 36 patients (subacute CVE), accounting for a total of 55 events (3.7%) between the procedure and 30 days. Event-free survival rates for CVE were 96.3% (95%CI: 95.3-97.3) at 30 days, 94.6% (95%CI: 93.3-95.7) at 1 year, and 92.9% (91.5-94.3) at 2 years. Acute CVE were disabling strokes in 79% of cases (**Table 3**). The risk of stroke peaked within the first 24 hours and levelled off within the first month, after which the risk of CVE remained stable over time (**Figure 3**). All-cause mortality during the observation period amounted to 32.6% (95%CI: 30-35.2%), and the occurrence of CVE within 30 days was associated with an increased risk of death (univariate HR [95%CI]: 2.62 [1.82-3.78], p<0.001) (**Figure 4**).

**Figure 2:**
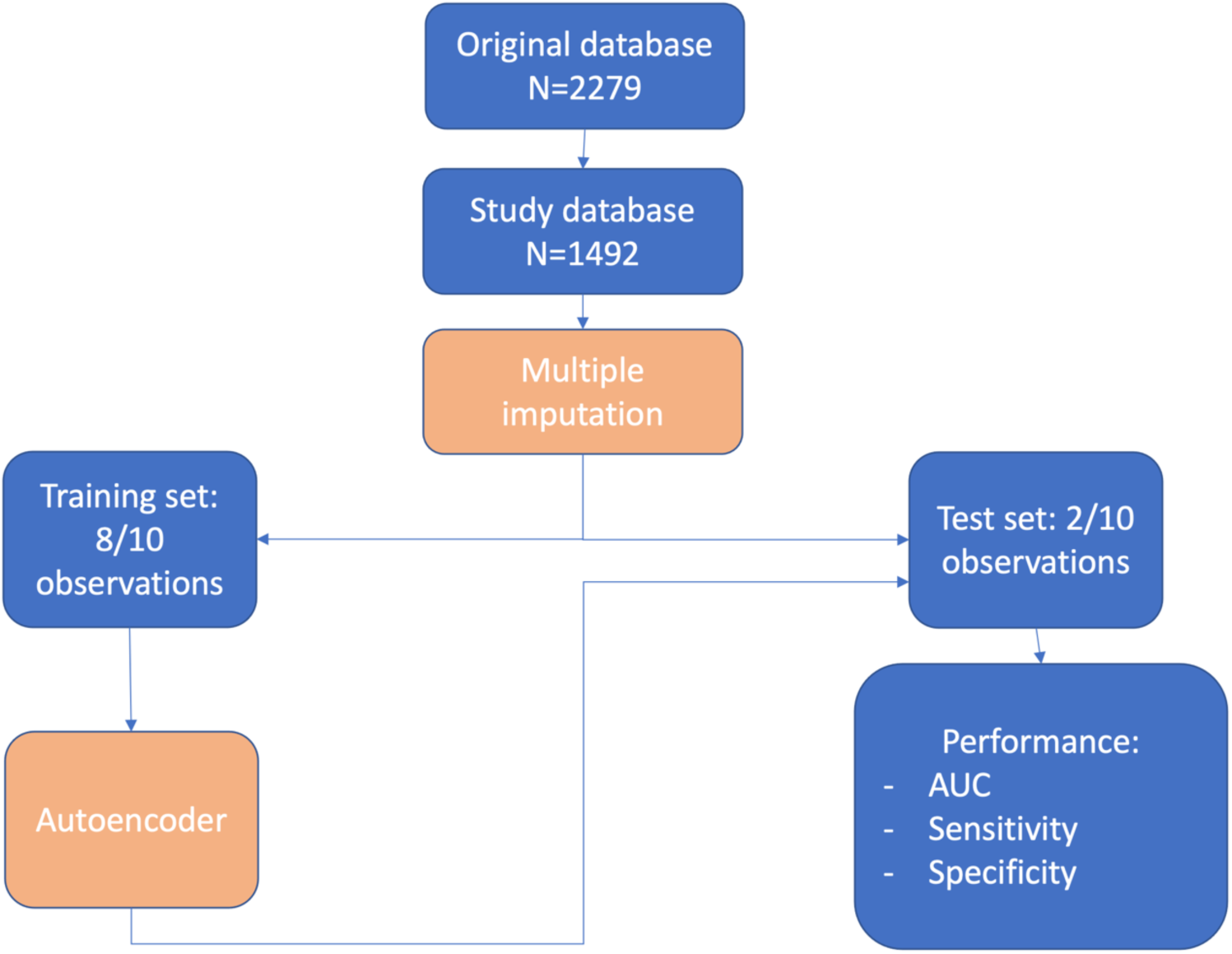
Study flow chart. HR: hazard ratio. MSCT: multi-slice computed tomography.

**Table 1.** Patient clinical characteristics by occurrence of cerebrovascular event within 30 days after TAVR.

|  | **Total**<br>**(n = 1492)** | **No CVE**<br>**(n=1437)** | **CVE**<br>**(n=55)** | **P value** |
| --- | --- | --- | --- | --- |
| Female, n (%) | 711 (47.7) | 685 (47.7) | 26 (47.3) | 0.99 |
| Age, years | 83.2 (79.4-86) | 83.1 (79.4-86.1) | 83.4 (79.3-87.4) | 0.27 |
| Height, cm | 165 (159-172) | 165 (159-172) | 165 (160-17) | 0.67 |
| Weight, kg | 71 (62-82) | 71 (62-82) | 67 (59-84.5) | 0.28 |
| STS-PROM (%) | 4.6 (3.2-7.0) | 4.6 (3.1-7.0) | 4.6 (3.1-7.0) | 0.45 |
| **Concomitant diseases/history** |  |  |  |  |
| Hypertension, n (%) | 1268 (85.0) | 1219 (84.8) | 49 (89.1) | 0.49 |
| Dyslipidemia, n (%) | 975 (65.3) | 941 (65.5) | 34 (61.8) | 0.67 |
| Diabetes mellitus, n (%) | 370 (24.8) | 363 (25.3) | 7 (12.7) | 0.051 |
| Dialysis, n (%) | 30 (2.0) | 29 (2.0) | 1 (1.8) | 0.99 |
| Chronic obstructive pulmonary disease, n (%) | 195 (13.1) | 187 (13) | 8 (14.5) | 0.89 |
| Coronary artery disease, n (%) | 945 (63.3) | 910 (63.3) | 35 (63.6) | 0.99 |
| History of cardiac surgery, n (%) | 224 (15.0) | 216 (15.0) | 8 (14.5) | 0.99 |
| Angina pectoris CCS 3 or 4, n (%) | 118 (7.9) | 113 (7.8) | 5 (9.1) | 0.54 |
| Prior syncope, n (%) | 183 (12.3) | 175 (12.2) | 8 (14.5) | 0.75 |
| Prior stroke/TIA, n (%) | 174 (11.7) | 162 (11.3) | 12 (21.8) | 0.029 |
| Prior carotid artery disease, n (%) | 168 (11.3) | 155 (10.8) | 13 (23.6) | 0.006 |
| Peripheral artery disease, n (%) | 220 (14.7) | 209 (14.5) | 11 (20) | 0.35 |
| Atrial fibrillation, n (%) | 492 (33) | 471 (32.8) | 21 (38.2) | 0.49 |
| Previous pacemaker implantation, n (%) | 139 (9.3) | 134 (9.3) | 5 (9.1) | 0.99 |
| **Medication at baseline** |  |  |  |  |
| Aspirin, n (%) | 929 (62.3) | 894 (62.2) | 35 (63.6) | 0.94 |
| Clopidogrel, n (%) | 273 (18.3) | 265 (18.4) | 8 (14.5) | 0.57 |
| Prasugrel, n (%) | 7 (0.5) | 7 (0.5) | 0 | 0.99 |
| Ticagrelor, n (%) | 30 (2) | 29 (2.0) | 1 (1.8) | 0.99 |
| Oral anticoagulation, n (%) | 431 (28.9) | 410 (28.5) | 21 (38.2) | 0.16 |
| Statin, n (%) | 784 (52.5) | 758 (52.7) | 26 (47.3) | 0.50 |
| **Laboratory values** |  |  |  |  |
| Creatinine, $\mu\text{mol/L}$ | 90 (74-112) | 90 (74-111) | 95 (79-120.5) | 0.128 |
| Haemoglobin, g/L | 122.50 (111-133) | 123 (111-133) | 118 (110-130) | 0.21 |
| Thrombocytes, $10^3/\text{mm}^3$ | 212 (172-259) | 212 (172-259) | 203 (173.5-252.50) | 0.72 |
| **Echocardiographic characteristics** |  |  |  |  |
| Mean aortic valve gradient, mmHg | 40 (29, 5) | 40 (29, 5) | 41 (33.5, 54) | 0.27 |
| Left ventricle ejection fraction, % | 60 (45, 65) | 60 (45, 65) | 60 (46.50, 65) | 0.36 |
| Mitral regurgitation grade III or IV, n (%) | 284 (19.2) | 270 (18.7) | 15 (27.3) | 0.46 |
| **Procedural characteristics** |  |  |  |  |
| **Main Access** |  |  |  |  |
| Transfemoral, n (%) | 1294 (86.7) | 1247 (86.8) | 47 (85.5) | 0.90 |
| Transapical, n (%) | 180 (12.1) | 173 (12.0) | 7 (12.7) |  |
| Other, n (%) | 18 (1.2) | 17 (1.2) | 1 (1.8) |  |
| **Concomitant procedure** |  |  |  |  |
| PCI, n (%) | 175 (11.7) | 168 (11.7) | 7 (12.7) | 0.98 |
| non-PCI interventions, n (%) | 68 (4.6) | 63 (4.4) | 5 (9.1) | 0.18 |
| **Type of valve** |  |  |  |  |
| Balloon-expandable, n (%) | 683 (45.8) | 657 (45.7) | 26 (47.3) | 0.96 |
| Self-expanding, n (%) | 688 (46.1) | 663 (46.1) | 25 (45.5) |  |
| Mechanically expandable, n (%) | 121 (8.1) | 117 (8.1) | 4 (7.3) |  |
| CCS = Canadian Cardiovascular Society grading; CVE = cerebrovascular event; PCI = percutaneous coronary intervention; STS-PROM = Society of Thoracic Surgeons Predicted Risk of Mortality; TIA = Transient ischemic attack.<br>Balloon-expandable valves: SAPIEN, SAPIEN XT, or SAPIEN 3 (Edwards Lifesciences, Irvine, CA, USA).<br>Self-expanding valves: CoreValve, Evolut R/PRO (Medtronic, Minneapolis, MN, USA), Portico (Abbott, Chicago, IL, USA), or Symetis ACURATE.ACURATE neo (Boston Scientific, Marlborough, MA, USA).<br>Mechanically-expanding valves: Lotus (Boston Scientific, Marlborough, MA, USA). |  |  |  |  |

**Table 2:** Patient CT imaging characteristics.

|  | **Total**<br>**(n = 1492)** | **No CVE**<br>**(n=1437)** | **CVE**<br>**(n = 55)** | **P value** |
| --- | --- | --- | --- | --- |
| Bicuspid valve on MDCT, n (%) | 398 (26.7) | 386 (26.8) | 12 (21.8) | 0.41 |
| Maximal annular diameter, mm | 26.8 (24.3-28.8) | 26.8 (24.2-28.8) | 27.3 (25.9-29.1) | 0.20 |
| Minimal annular diameter, mm | 21.3 (19.7-23.6) | 21.4 (19.7-23.6) | 21 (19.2-22.9) | 0.28 |
| Mean annular diameter, mm | 23.7 (21.9-25.4) | 23.7 (21.9-25.4) | 23.8 (22.6-25.3) | 0.30 |
| Annulus area, cm2 | 431.4 (352.5-496.6) | 431.4 (351.6-496.2) | 441.2 (384-503.8) | 0.28 |
| Annulus perimeter, mm | 77.8 (73.3-85.2) | 77.8 (73.3-85.2) | 78.1 (74.2-84.6) | 0.95 |
| Annulus eccentricity | 0.78 (0.73-0.84) | 0.78 (0.73-0.84) | 0.76 (0.71-0.81) | 0.052 |
| Left coronary artery height, mm | 15.3 (12.9-18.8) | 15.3 (12.9-18.5) | 15.1 (12.9-17.9) | 0.86 |
| Right coronary artery height, mm | 16.9 (14.7-19.3) | 16.9 (14.6-19.3) | 17 (15.6-18.9) | 0.75 |
| Ascending aorta diameter, mm | 32.3 (29.4-34.9) | 32.3 (29.9-34.9) | 31.5 (29.8-34.5) | 0.73 |
| Ascending aorta length, mm | 63.8 (57.0-69.1) | 63.8 (57.0-69.1) | 62.6 (58.1-69.1) | 0.91 |
| Sino-tubular junction diameter, mm | 28.6 (26.3-31.1) | 28.6 (26.3-31.2) | 28.3 (26.0-30.7) | 0.55 |
| Total aortic valve calcium volume, mm3 | 176.1 (42.5-415) | 176.1 (40.4-410.7) | 204.2 (99.4-453.5) | 0.23 |
| Left cusp aortic valve calcium volume, mm3 | 61.8 (18.3-167.6) | 61.6 (18.2-167.8) | 67.1 (26.4-161) |  |
| Right cusp aortic valve calcium volume, mm3 | 53.5 (15.3-134.5) | 53 (14.9-131.5) | 95.6 (26.7-174.5) | 0.04 |
| Non-coronary cusp aortic valve calcium, mm3 | 86.5 (28.6-203.9) | 86 (28-203.6) | 118.2 (40.9-249.4) | 0.20 |
| LVOT calcium, total, mm3 | 0.20 (0.0-33.4) | 0.20 (0.0-33.9) | 0.10 (0.0-26.8) | 0.84 |
| Left cusp LVOT calcium, mm3 | 0.0 (0.0-3.3) | 0.0 (0.0-3.2) | 0.0 (0.0-5.6) | 0.28 |
| Right cusp LVOT calcium, mm3 | 0.0 (0.0-0.0) | 0.0 (0.0-0.0) | 0.0 (0.0-0.1) | 0.034 |
| Non-coronary cusp LVOT calcium, mm3 | 0.0 (0.0-0.0) | 0.0 (0.0-0.0) | 0.0 (0.0-0.3) | 0.52 |
| Mitral valve total calcium volume, mm3 | 11.9 (0.0-321.9) | 11.7 (0.0-308.3) | 35.6 (0.0-662.4) | 0.17 |
| Mitral anterior leaflet calcium volume, mm3 | 0.50 (0.0-56.6) | 0.50 (0.0-56.4) | 1.80 (0.0-80.3) | 0.26 |
| Mitral posterior leaflet calcium volume, mm3 | 17.8 (0.0-278.3) | 16.9 (0.0-274.3) | 34.3 (0.0-649.1) | 0.24 |
| LVOT = left ventricular outflow tract. |  |  |  |  |

**Table 3.** Incidence of cerebrovascular event.

|  | Day 0 | Day 1 to 30 days | > 30 days | Total CVE |
| --- | --- | --- | --- | --- |
| CVE, n (%) | 19 (1.3) | 36 (2.4) | 61 (4.1) | 105 (7.8) |
| Disabling stroke, n (%) | 15 (1.0) | 16 (1.1) | 31 (2.1) | 62 (4.2) |
| Non-disabling stroke, n (%) | 2 (0.1) | 13 (0.9) | 15 (1.0) | 30 (2.0) |
| TIA, n (%) | 2 (0.1) | 7 (0.5) | 15 (1.0) | 24 (1.6) |
| CVE = Cerebrovascular event; TIA = Transient ischemic attack. |  |  |  |  |

**Figure 3:**
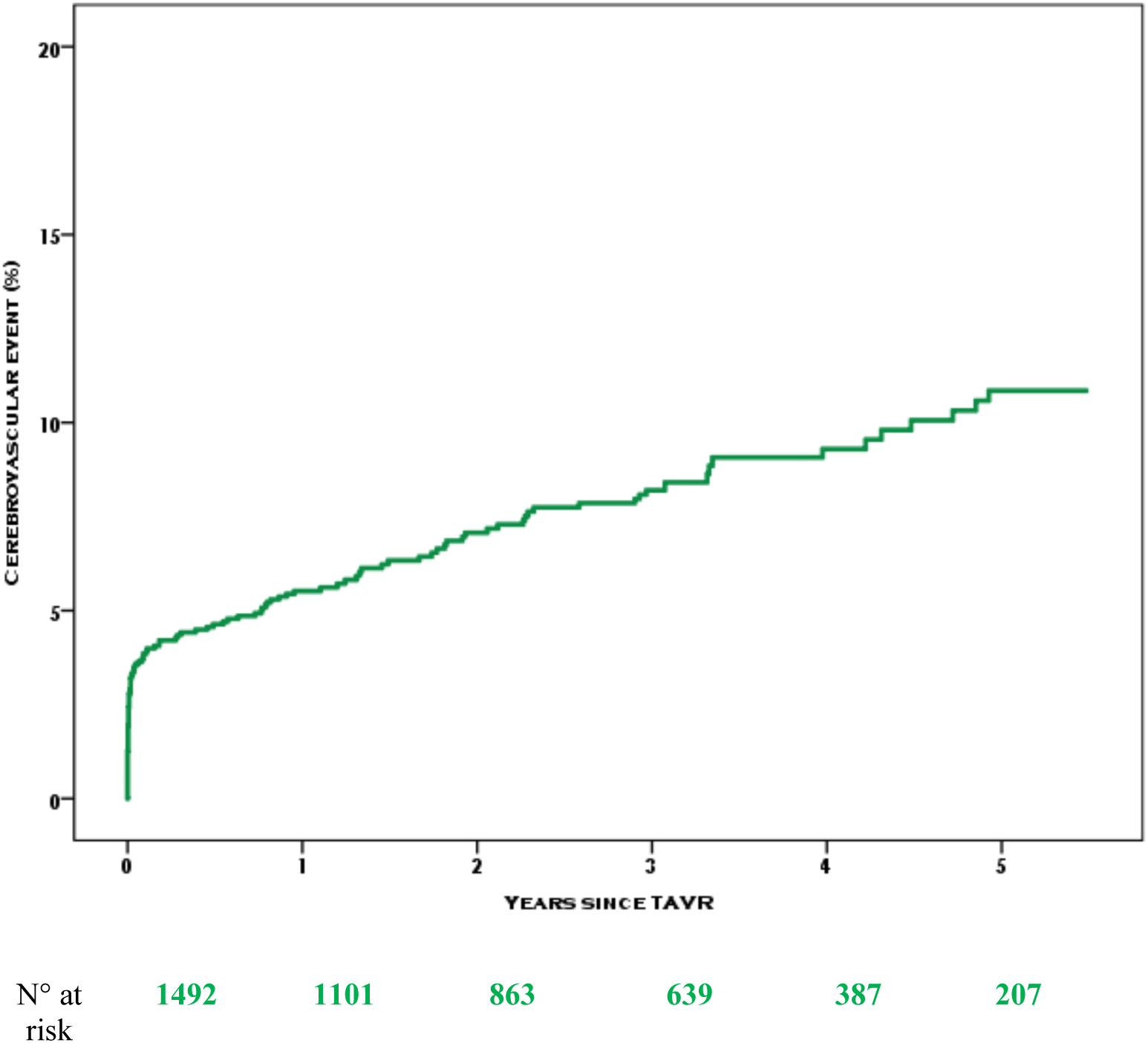
Kaplan-Meier curve of cerebrovascular events after TAVR.

**Figure 4:**
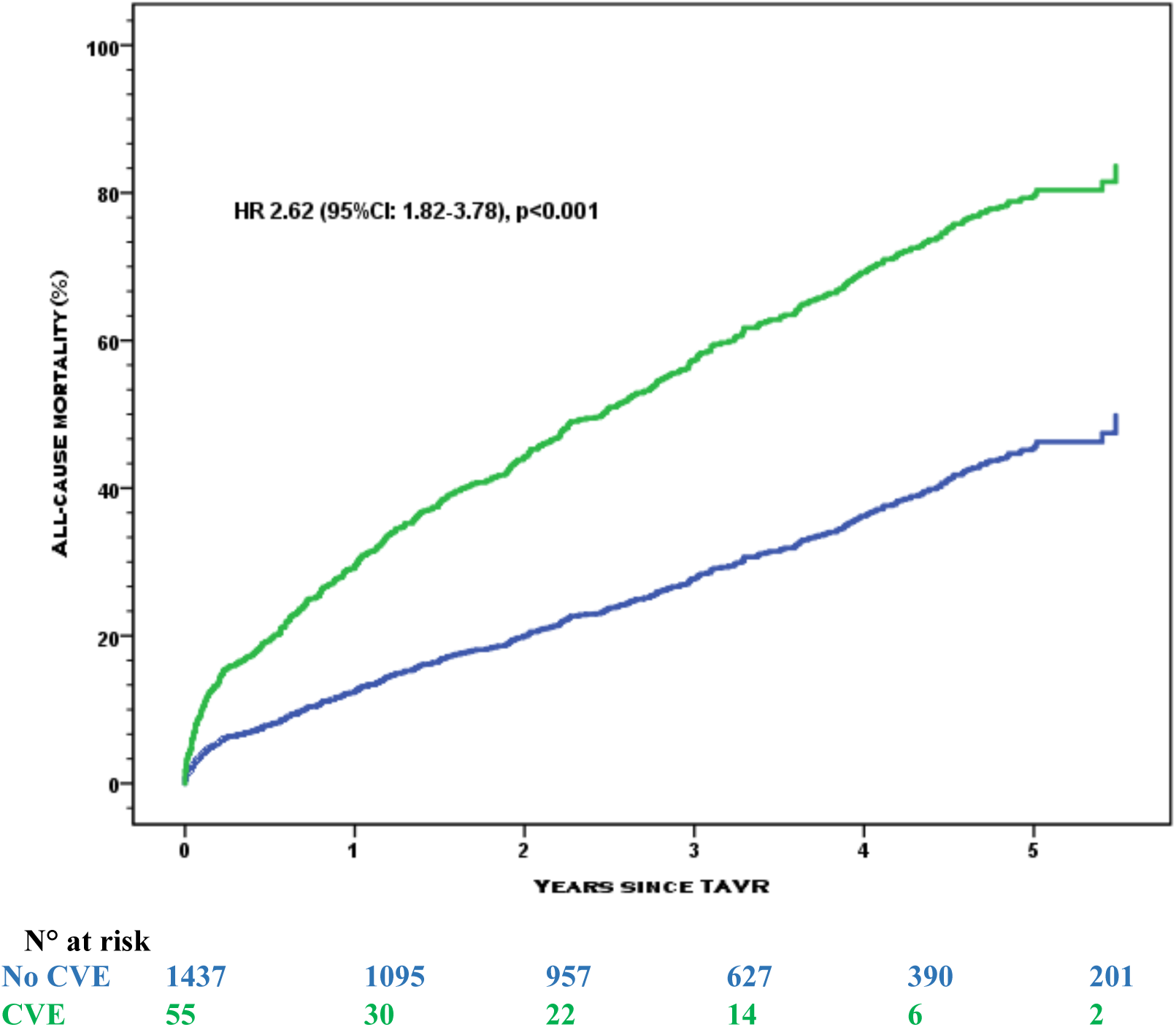
Kaplan-Meier curve of all-cause mortality separated by occurrence of cerebrovascular event within 30 days of TAVR.

### TAVR-related CVE predictive model

An extensive set (>100 variables) of clinical, biological, imaging, and procedural characteristics as well as complications were considered for the deep learning predictive modelling (**Tables 1, 2**). Recursive feature elimination by variable importance suggested a drop in predictive performance when less than the 63 variables were included in the model (**Supplementary Figure 2**). The trained model included 2 hidden layers in the encoder and decoder and used MDCT data (variables represented aortic calcium volume, aortic annulus and left ventricle outflow tract dimensions, ascending aorta dimensions and angulation), along with other clinical data (**Figure 1, Supplementary Figure 2, Supplementary Tables 1 and 2**). The constructed predictive model had an AUC of 0.79 (0.65-0.93) (**Figure 5**).

**Figure 5:**
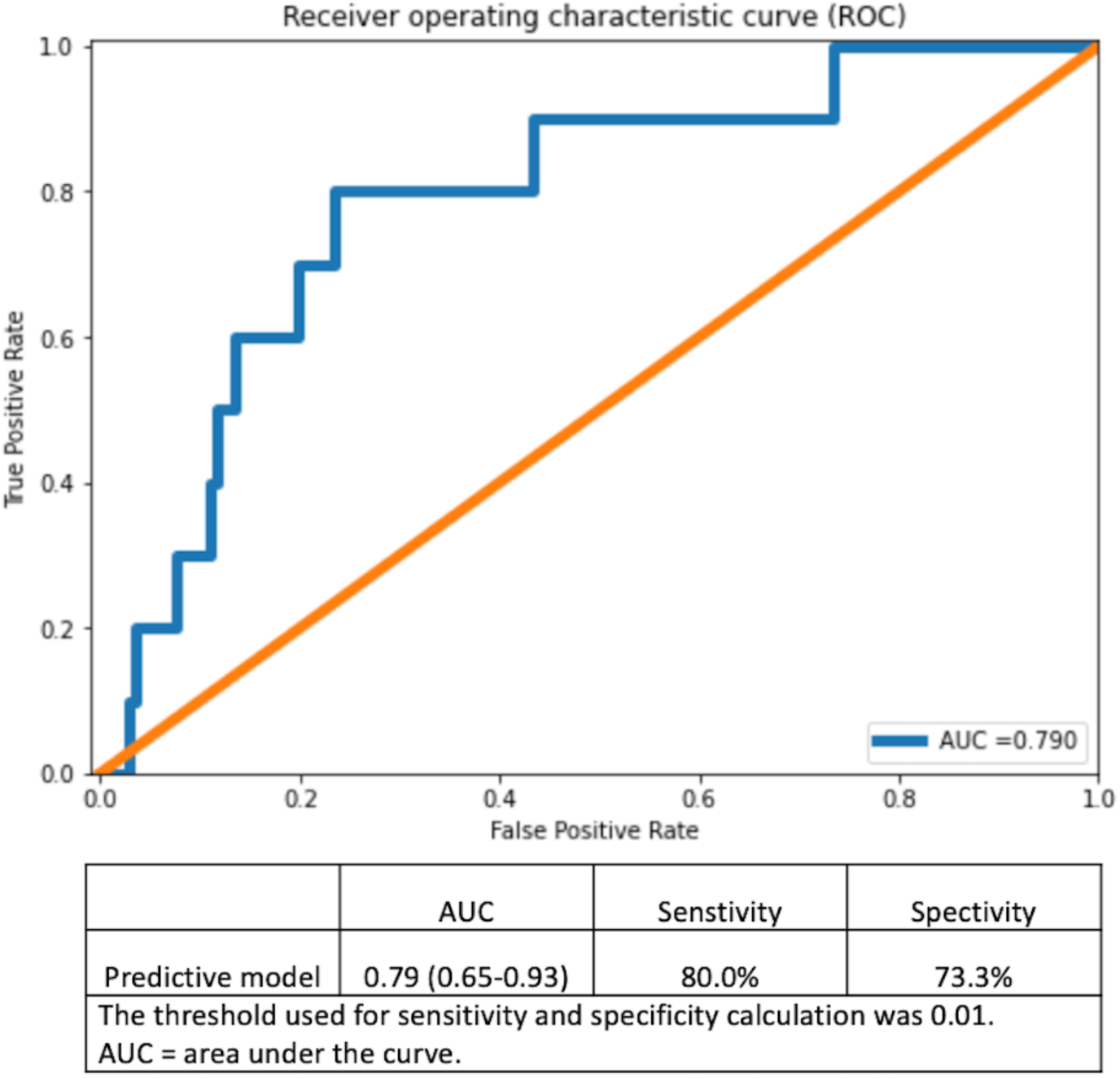
Receiver operating curve of the predictive model. The mean squared error between predicted and known values on the test set was 0.01.

## DISCUSSION

Herein we propose a novel predictive model for TAVR-related CVE. The proposed model implements the deep neural network-type of algorithm and yields a satisfactory predictive performance with an AUC of 0.79 (0.65 to 0.93). Our study also suggests that imaging data could be as important as clinical characteristics for the construction of predictive models for cardiovascular procedures, although both types of variables are used in synergy by the constructed model developed for CVE.

CVE remains one of the most feared adverse events complicating TAVR. Despite fifteen years of improvement of the technique and refinement of TAVR devices, the occurrence of CVE during the peri-operative period remains notable^10,11^. Preventive strategies for TAVR-related CVE are yet to provide evidence for their effectiveness. Cerebral protection devices have so far not been proven to reduce the risk of clinical CVE and none of the investigated antithrombotic strategies has shown protective effects in clinical investigations^26-29^. The need for further investigation has been underscored by the Academic Research Consortium Initiative consensus proposing a standardized neurological endpoint evaluation for clinical research (NeuroARC)^23^.

Previous studies have identified various risk factors for CVE in patients undergoing TAVR, which included clinical data such as atrial fibrillation and history of CVE, imaging data such as calcification of the aortic valve complex and reduced native and prosthetic aortic valve area, and procedural data such as balloon dilatation and device dislocation or embolization^9,13-18^, however, a prediction model for stroke integrating all these factors have not been developed to date. Our data suggest an intricate relationship between clinical characteristics, anatomical features, peri-interventional antithrombotic management, and procedural complications to yield an overall risk estimation for CVE^30,31^. Albeit severe, CVE after TAVR is an infrequent event which renders the classical predictive approach based on univariate then multivariable regression techniques difficult to implement. The multifactorial nature of peri-TAVR CVE makes this approach further difficult. As suggested in our study, a justified deep learning approach has the potential to overcome such difficulties. Although, imaging parameters have not been identified as associated with CVE on conventional univariate analysis (**Table 2**), the autoencoder model exploited them to yield a satisfactory predictive performance. More interestingly, we observed a synergic exploitation of imaging data along clinical characteristics which suggests that predictive modelling could benefit from the inclusion of diverse types of data for their complementarity. This is, to the best of our knowledge, the first study proposing a predictive tool for TAVR-related CVE. The constructed model has been implemented by means of an open source online calculator (https://www.welcome.alviss.ai/#/cvecalculator). This online calculator may help improve risk stratification of patients undergoing TAVR and tailor subsequent follow-up and management strategies by recognizing high risk of CVE.

Precision medicine aims at adapting decision to a patient given his/her characteristics intended for a deep phenotyping. Machine learning models allow a holistic approach by analysing complex interactions between an extensive number of patient characteristics. In the present investigation, medical history, symptoms, treatment, imaging and procedural characteristics were entered and analysed using an autoencoder algorithm. Our results suggest an existing ground for the interaction between imaging features such as valve tissue calcification and dimensions of the aortic valve complex, and clinical features to yield CVE. Additional imaging data such as MRI, extensive biological (including per-operative coagulation function estimation or genetics) and environmental profiling may further improve risk assessment. However, the predictive model would require to be adapted for routine clinical practice. The present study is another example of an increasing implementation of machine learning tools to analyse large healthcare databases^32^.

### Limitations

First, the Bern TAVR registry is a manually built database prone to transcription error although our results are consistent with previous reports^33,34^. Second, frailty indexes are lacking in the present registry and could improve the predictive performance. Third, given constraints relative to our model we could provide only relative variable weighting to explain its functioning^4^. Forth, CT imaging characteristics were obtained using the 3mensio software, however any other software using 3D reconstruction would allow operators to obtain the measurements. Fifth, the cohort studied in the present study is representative of a Western population in a single-center. As the predictors of CVE might differ in a different population because of ethnic, environmental, and genetic factors, that were not recorded in the registry, further external validation in an independent cohort is warranted. Lastly, this is an observational study based on a large prospective TAVR registry including nearly 1,500 patients. The occurrence of TAVR-related CVE was however observed in 55 patients only. We used the rare event autoencoder for prediction to address this limitation, however, the small number of events did not allow separate analyses of acute (procedural) and subacute CVE, respectively. Future machine learning research with a larger population is warranted to develop a refined model for predicting TAVR-related CVE.

## CONCLUSIONS

Despite their complex pathophysiology and rarity, TAVR-related CVE can be predicted by using artificial neural networks. The model was implemented online for broad usage (https://www.welcome.alviss.ai/#/cvecalculator). The proposed approach illustrates the potential of artificial intelligence in developing prediction models for multifactorial adverse events such as CVE.

## Data Availability

The data underlying this article is held at the Clinical Trials Unit of the University of Bern, Switzerland. Data can be made available to external investigators upon request to the corresponding author with permission of the institution, according to local regulations.

## CONTRIBUTORS

PO, TP conceived the study. PO, TP had responsibility for the design of the study. PO, TP, TO, MA, DJ, SS, FP, JL, GS, CG, SW were responsible for the acquisition of data. PO did the analysis and interpreted the results in collaboration with TP, SW and all other authors. TO, PO, TP wrote the first draft of the report. All authors critically revised the report for important intellectual content and approved the final version.

